# Early Stance Negative Knee Joint Powers and Knee Flexion Moments Can Be Tracked Using Inertial Measurement Units

**DOI:** 10.1101/2025.10.17.25338247

**Authors:** Fany Alvarado, Julien A. Mihy, Mayumi Wagatsuma, Millissia A. Murro, Jocelyn F. Hafer

**Affiliations:** University of Delaware

**Keywords:** angular velocity, gait, wearable sensors, kinetics

## Abstract

Joint kinetics (e.g., moments and powers) are important markers of mobility and joint health but their measurement is generally limited to lab settings. However, we may be able to estimate kinetics with portable inertial measurement units (IMUs) by detecting important movement features that contribute to or occur simultaneously with kinetics. This study aimed to investigate whether early stance segment and joint angular velocities measured with IMUs correlate with early stance negative knee joint power and knee flexion moments. We collected motion capture, force plate, and IMU data during gait for healthy young adults, older adults, and older adults with knee OA. Peak knee flexion moments and early stance negative knee joint powers were calculated from motion capture data using inverse dynamics. Segment (thigh and shank) and joint (knee) angular velocities were calculated from IMU gyroscope data, and peaks in the first 5-25% of stance were identified. Pearson correlation coefficients were evaluated between motion capture data (peak knee flexion moments and negative knee joint powers) and IMU data (peak thigh, shank, and knee angular velocities). Significant correlations were found between negative knee joint powers and shank and knee joint angular velocities (r=0.59, p<0.001; r=- 0.37, p= 0.004, respectively). Knee flexion moments significantly correlated with shank angular velocities (r=-0.44, p<0.001). Our results suggest the potential to track longitudinal changes in knee joint power outside the lab with only a shank sensor.

## 1. Introduction

Knee joint kinetics during gait are important markers of mobility and joint health. For example, adults with knee osteoarthritis (OA) often have smaller peak knee flexion moments [1], [2] and larger knee adduction moments [3], [4] compared to asymptomatic adults. Such differences in kinetics have been associated with OA severity and progression [5], [6]. Because of the relationship between knee kinetics and disease status, some interventions aim to alter knee kinetics in adults with or at risk of knee OA. However, the application and evaluation of these interventions beyond lab settings are limited because measurement of joint kinetics generally requires optical motion capture and force plates (MoCap).

Previous studies estimated or predicted joint kinetics without MoCap using wearable sensors in combination with musculoskeletal modeling [7], [8] or machine learning [9], [10]. While such approaches have successfully estimated kinetics, they are limited to research teams with specialized expertise or to patient groups where appropriate model training data are accessible. An alternative and potentially more accessible approach to assessing kinetics without MoCap may be to capture surrogate metrics that can detect differences or changes in kinetics without advanced computational methodology. We previously demonstrated that angular velocities captured with inertial measurement units (IMUs) can track propulsive hip and ankle joint powers [11]. A similar approach may provide surrogate metrics for kinetics that are important in knee OA.

The knee joint kinetics that are linked to knee OA status or progression – knee flexion and adduction moments calculated from inverse dynamics – occur early during the stance phase of gait. While IMUs do not directly measure the position or force data necessary to directly estimate joint moments, they accurately measure angular velocity, which is a component of joint power. Joint powers are less commonly reported in knee OA literature compared to joint moments, but negative knee joint power during early stance may be a meaningful marker of joint health. Negative knee joint power occurs around the same time as knee flexion and adduction moments and represents the energy absorbed by the knee during loading. A decrease in knee joint power has been associated with decreased quadriceps strength [12]. Individuals with knee OA have decreased quadriceps muscle strength compared to asymptomatic adults [13] and older adults also have lower quadriceps muscle strength [14] and negative knee joint powers [15] compared to younger adults. Negative knee joint power and knee flexion moments are influenced similarly by gait modifications like altered stride length, cadence, or speed [16], [17], [18] with increases in joint powers and moments as stride length or speed increase. This suggests that negative knee joint power may track with meaningful changes in other knee kinetics, such as those that may be targeted in interventions for reducing the declines caused by aging or knee OA. Thus, it would be useful to develop methods that would enable us to track longitudinal changes in kinetics.

In this study, we sought to investigate whether early stance segment and joint angular velocities measured by IMUs correlate with early stance negative knee joint power and knee flexion moments. We included participants from different populations (younger healthy, older asymptomatic, older with knee OA) in order to test whether this correlation held across a spectrum of expected variance in knee kinetics. We hypothesized that knee, thigh, and shank angular velocities would correlate with negative knee joint power and knee flexion moments. Our primary analysis examined these correlations across all participant groups combined and we performed additional analyses to explore whether the correlations within each group were similar to the correlation across all three groups.

## 2. Methods

### 2.1 Participants

Ten healthy younger adults (25.4±2.5 yrs), 10 healthy older adults (61.2±4.6 yrs), and 10 older adults who self-reported a previous diagnosis of knee OA by a physician (64.2±2.9 yrs) were recruited via word of mouth, flyers, and a recruitment email (Table I). Participants were eligible to participate if they were able to walk for 30 minutes without any assistive devices and had no history of major injury limiting physical activity, surgery, or chronic pain in the lower extremities or back (except for the knee in the knee OA group). Exclusion criteria included neurological disorders or a history of stroke, a cardiovascular or pulmonary condition that limits daily activity, uncontrolled diabetes, vertigo, or dizziness. Participants in the knee OA group were excluded if they had a joint injection in the last 3 months before participating. Before completing any study procedures, all participants completed an informed consent that was approved by the university’s institutional review board (IRB: 1846956-1).

**Table 1.** Participant characteristics.

|  | Old |  | OA |  | Young |  |
| --- | --- | --- | --- | --- | --- | --- |
|  | Mean | SD | Mean | SD | Mean | SD |
| n | 10 |  | 10 |  | 10 |  |
| Age (yrs) | 61.2 | 4.6 | 64.2 | 2.9 | 25.4 | 2.5 |
| Height (cm) | 172.1 | 13.5 | 168.2 | 12.9 | 167.7 | 9.0 |
| Body Mass (kg) | 71.5 | 20.8 | 70.8 | 13.2 | 72.4 | 11.6 |
| Preferred Speed (m/s) | 1.5 | 0.2 | 1.4 | 0.1 | 1.4 | 0.2 |
| Fast Speed (m/s) | 2.0 | 0.2 | 1.8 | 0.2 | 1.9 | 0.2 |
| KOOS | 96.3 | 5.0 | 75.3 | 12.2 | 98.5 | 2.0 |

Each group included an equal number of females and males.
Osteoarthritis (OA)

### 2.2 Data Collection

All participants filled out the Knee Injury and Osteoarthritis Outcome Score (KOOS) to document knee-related symptoms and function. Demographics, weight, and height were collected. We then determined participants’ preferred walking speed as they walked across a 6m walkway that included space for acceleration and deceleration [19]. The average speed across five walking trials collected with timing gates was used as the preferred speed for data collection. Markers were placed according to a six-degree-of-freedom lower extremity marker set with anatomical markers placed on the posterior and anterior iliac spines, greater trochanter, medial and lateral femoral epicondyles, medial and lateral malleoli, 1st and 5th metatarsal heads, and medial and lateral heel. Rigid clusters were used to track the thigh and shank and the foot and pelvis were tracked using their anatomical markers. Participants wore 7 IMUs (APDM Opal v2, Clario, Philadelphia, USA). The pelvis sensor was placed on the sacrum, thigh and shank sensors were placed at the midpoint of each segment on the lateral aspect (under marker clusters), and the foot sensor was placed in a pouch and laced to the dorsum of the shoe.

Motion Capture (MoCap) and IMU data were collected simultaneously. MoCap data were collected using a 12-camera motion analysis system (Qualisys, Göteborg, Sweden) with three force plates (AMTI, Watertown, USA). Force plate and marker data were collected at 1200Hz and 120Hz, respectively. IMUs collected 3-axis accelerometer, gyroscope, and magnetometer data at 128 Hz in synchronized logging mode. After marker and IMU outfitting, participants performed calibration trials for MoCap (static standing) and IMU (static standing, straight line walking, and toe touches). Each participant then completed ten successful walking trials (5 per right and left side with clean force plate contact) within ±5% of their pre-determined preferred speed.

Then, participants walked at a self-selected fast speed (instructions: “walk as if you were about to catch a bus”) until they had completed ten successful walking trials that fell within a speed range of ±5%.

## 3. Analysis

### 3.1 IMU Analysis

Raw IMU data were collected in a reference frame oriented relative to the sensor housing. These data were then oriented to a world reference frame and a functional frame.

World orientation: IMU accelerometer, magnetometer, and gyroscope data were oriented to a world reference frame using a standard Kalman filter provided by the sensor manufacturer (APDM). For world orientation, the vertical (Z) axis is aligned with gravity and X and Y axes are orthogonal in the horizontal plane.

Functional orientation: Static standing, straight-line walking, and toe touches were used to orient IMU data to functional segment reference frames [20] (Fig.1). Static standing was identified as a period when the pelvis raw linear acceleration (gravity removed) and angular velocity signals were stable and around 0 magnitude. Each sensor’s average acceleration vector during this static standing period defined the functionally oriented inferior-superior axis. Functional movements (walking or toe-touches) were used to identify segment medial-lateral axes. Straight-line walking was used to identify the medial-later functional axis for the thigh, shank, and foot and toe-touches were used to identify the medial-lateral functional axis for the pelvis using principal component analysis (PCA). Each segment’s anterior-posterior axis was created as the cross-product of the inferior-superior (Z) and medial-lateral (X) axes. To create orthogonal reference frames, the inferior-superior axis (Z) was re-oriented by crossing the medial-lateral (X) and anterior-posterior (Y) axes.

**Fig. 1.**
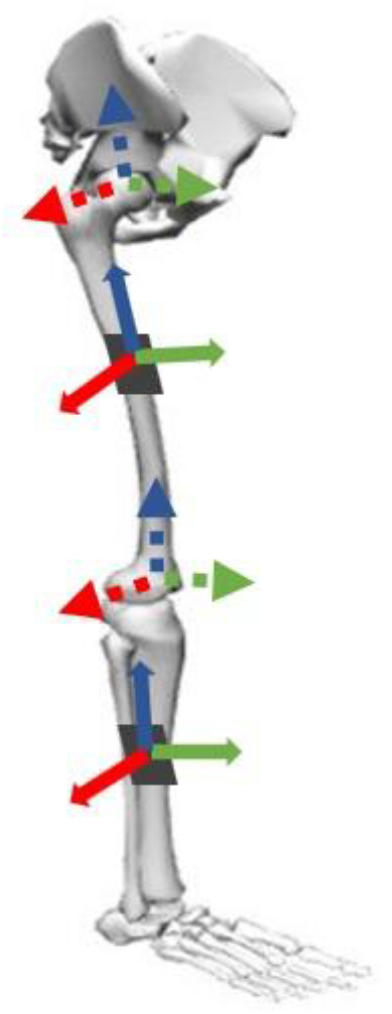
Illustration of IMU- and MoCap-based reference frames. Orientation of IMU functional reference frames illustrated with solid arrows. Location and orientation of MoCap anatomical reference frames illustrated with dotted arrows. Anterior-posterior axes (Y) are shown in green, medial-lateral axes (X) in red, and longitudinal axes (Z) in blue. Dark squares represent IMUs on the lateral thigh and shank.

Walking identification: Walking bouts were identified based on repeated oscillations of the shank sensor. Five second windows of the shank sensor’s raw angular velocity data were passed through a Fast Fourier Transform (FFT) and the frequency power from all three axes was summed to determine the total power density within each window. Windows with peaks in frequency power between 0.5 and 2.2 Hz were considered walking. Consecutive windows identified as walking were combined.

Gait events: We used data from foot sensors to identify gait events in each bout. World frame foot sensor Z axis (i.e., vertical) linear acceleration data were passed through a one-dimensional continuous wavelet transform (CWT). The first (highest-frequency) wavelet was low pass filtered with a second-order bidirectional (filtfilt MATLAB function) Butterworth filter at 4Hz. Gait events were identified when signal peaks were above half the median magnitude of all peaks in the filtered signal. We used a zero-velocity update (ZUPT) algorithm [20] (thresholds: <1 m/s2 acceleration magnitude after gravity removal, <1 rad/s angular velocity magnitude) to determine when the foot was static. We integrated world frame linear acceleration between static foot timepoints to get linear velocity and linear displacement. Integration from acceleration to velocity was drift corrected, so that the end and beginning were set to 0. The slope of vertical displacement was used to determine whether each identified gait event was a heel strike or toe-off. Gait events that occurred at a time when the slope of vertical foot displacement was negative were heel strikes, and gait events that occurred at a time when the slope of vertical foot displacement was positive were toe offs. Strides were created when consecutive gait events included heel strike-toe off-heel strike patterns.

Outcome variable calculation: Note that, because the focus of this study was on accessible surrogate metrics and not on direct estimates or predictions of joint kinetics, we did not attempt to replicate MoCap methods with IMU data. IMU outcome variables were derived from the functional medial-lateral axis of the thigh and the shank segments’ angular velocities. Knee angular velocity was determined by subtracting shank angular velocity from thigh angular velocity (we previously showed that this approach accurately replicates sagittal knee joint excursion [21]). IMU bouts that matched successful MoCap bouts were included for each speed. All strides were time normalized to 100% stance for the IMU data. Peak angular velocities during early stance (5-25% of stance phase) and times when peaks occurred (% stance) were identified for the thigh (local maximum), shank (local minimum), and knee (local maximum) (Fig.2). Only data for steady state strides were included in the final analysis. Steady state strides were those that differed by less than 0.1 m/s for stride speed and deviated less than 10 degrees from the horizontal direction compared to the prior or following stride. Outcome variables were averaged per speed for each subject.

**Fig. 2.**
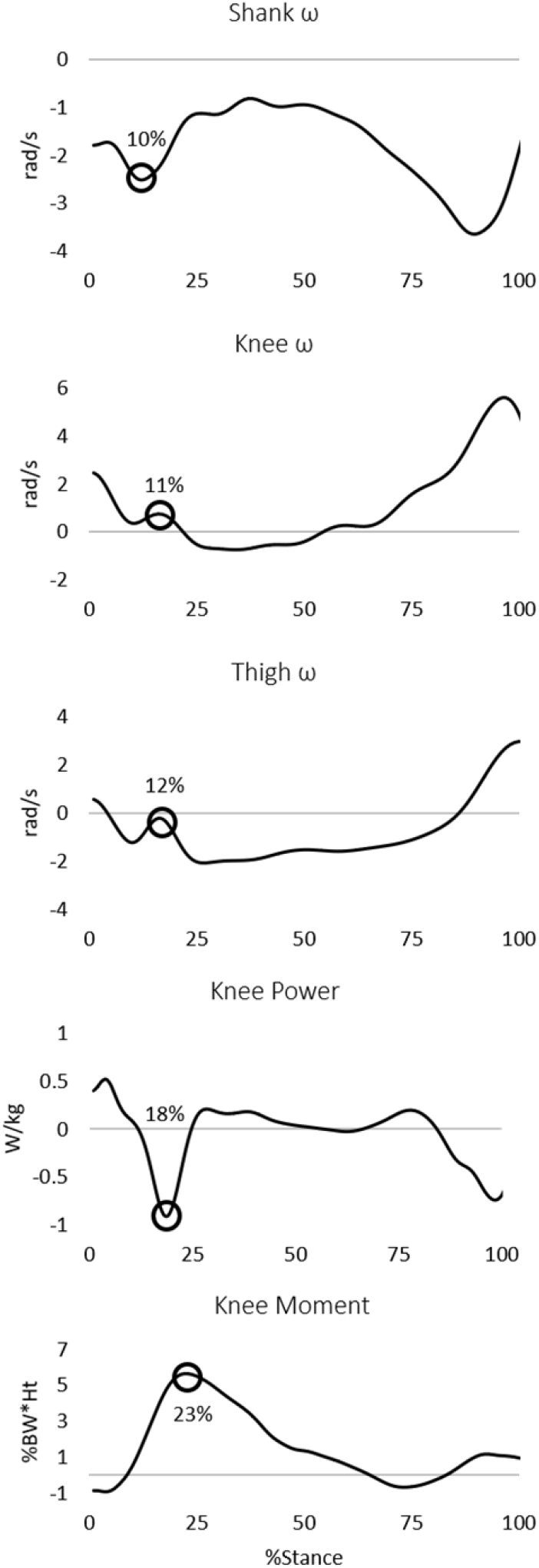
Segment and joint angular velocities during stance were estimated from functionally oriented IMU data, and stance phase sagittal knee powers and moments were calculated via inverse dynamics. Detected peaks of interest were calculated in early stance (5-25%), and their average timing during the gait cycle is indicated with a circle.

### 3.2 Motion Capture Analysis

Pre-processing: All MoCap data were imported into Visual 3D (HAS Motion, Kingston, Canada). The static standing trial was used to define the anatomical reference frame for each segment. Force plate and marker data were low pass filtered at 15Hz and 8Hz respectively.

Gait events: Automatic gait events were determined using ground reaction forces. Heel strike was the first frame when the ground reaction force exceeded 20N and toe-off was the final frame with a ground reaction force >20N.

Outcome variables: External knee joint moments and knee joint powers were calculated using standard inverse dynamics procedures [22], [23] and reported in the thigh reference frame. All kinetics were time normalized to 101 points per stance phase (100% stance). Knee joint powers were normalized to body mass (i.e., W/kg) and moments were normalized to body weight and height (i.e., %BW*Ht). Stance phase power and moment times series data were exported to MATLAB, where early stance negative power and peak knee flexion moment magnitudes and timings (% stance) were identified. Outcome variables were averaged per speed for each subject.

### 3.3 Statistical Analysis

We tested whether MoCap kinetics could be tracked using IMU-derived angular velocities using Pearson correlation coefficients. Correlations were performed between MoCap data (peaks in early stance negative knee joint power and knee flexion moments) and IMU data (peaks in medial-lateral axis thigh, shank, and knee angular velocities) across the two different speeds. All data were included for analysis, even those identified as potential outliers, which were assessed using a standard boxplot method. Exploratory correlations were performed across all three groups between MoCap data and IMU-derived angular velocities.

## 4. Results

Early stance negative knee joint power correlated with shank segment and knee joint angular velocities (r=0.60, p<0.001; r=-0.37, p=0.005, respectively; Fig. 3), but this correlation did not reach significance for the thigh segment angular velocity (r=-0.06, p=0.655). Peak knee flexion moment was correlated with shank angular velocity (r=-0.44, p<0.001), but not knee joint and thigh segment angular velocities (r=0.20, p=0.124; r=-0.01, p=0.927). Peak knee joint negative power and flexion moment occurred at 17.5±1.8% and 23.2±2.4% of stance on average. Peaks for knee joint angular velocity and thigh and shank segment angular velocities occurred at 11.2±0.5%, 11.2±1.9%, and 10.3±3.4% of stance on average. Our exploratory analysis of correlations within each group largely agreed with our primary findings (Fig. 4 and 5). Early stance negative knee joint power was correlated with shank segment angular velocity in the OA (r=0.81, p<0.001) and young (r=0.78, p<0.001) groups, but not in the older group (r=0.42, p=0.064). Peak knee joint power correlated with knee joint angular velocity only in the young group (r=-0.61, p=0.004), but not in the OA or older groups (r=-0.33, p=0.154; r=-0.31, p=0.182).

**Fig. 3.**
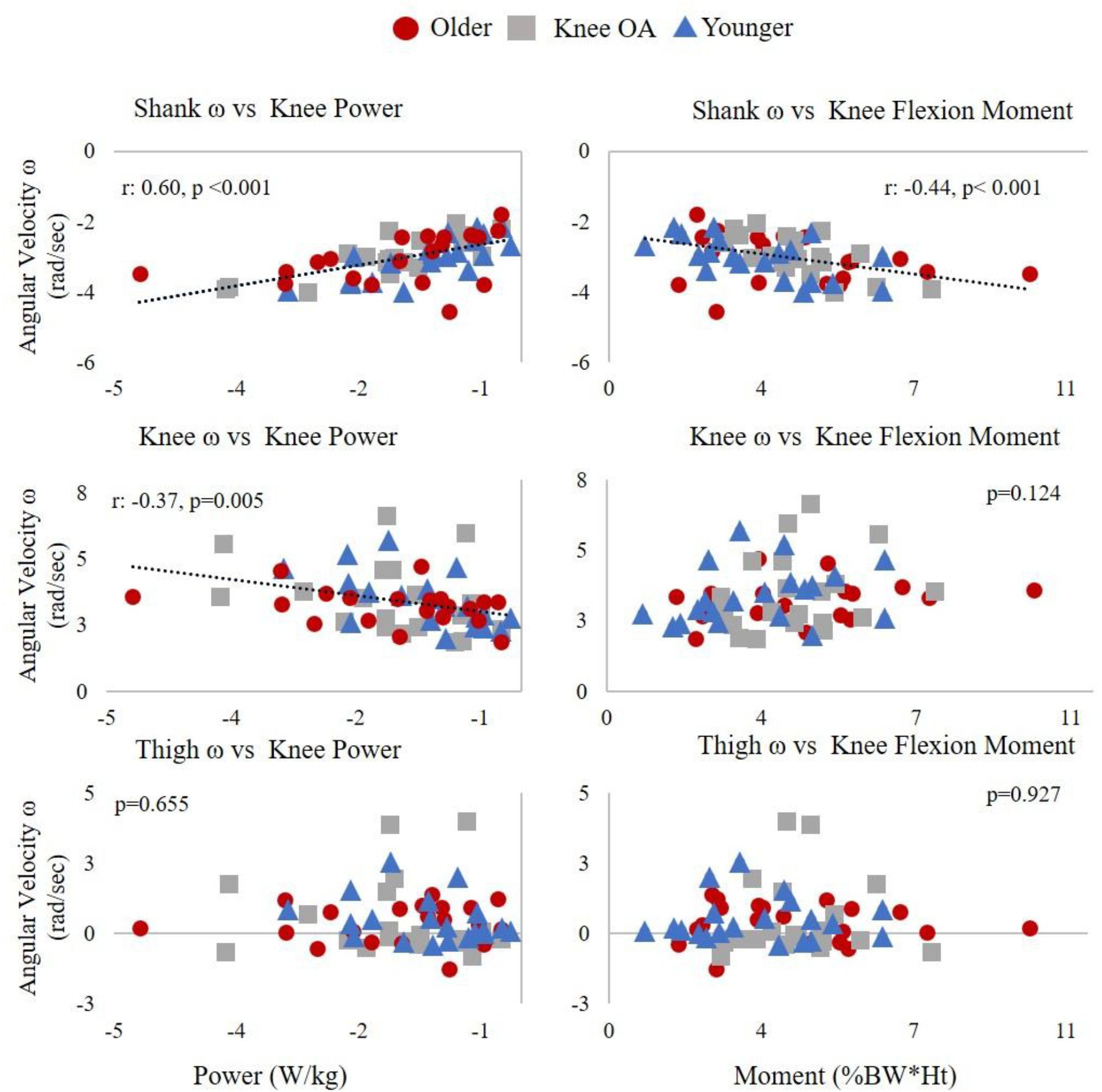
Early stance segment angular velocities measured with IMUs are plotted against early stance negative knee joint power and knee flexion moments calculated with inverse dynamics across all groups. Each data point represents an individual participant’s average value during preferred or fast speed walking. P-values are shown for each comparison and trend lines and Pearson correlation coefficients are included where correlations were significant. All r values can be found in the text.

**Fig. 4.**
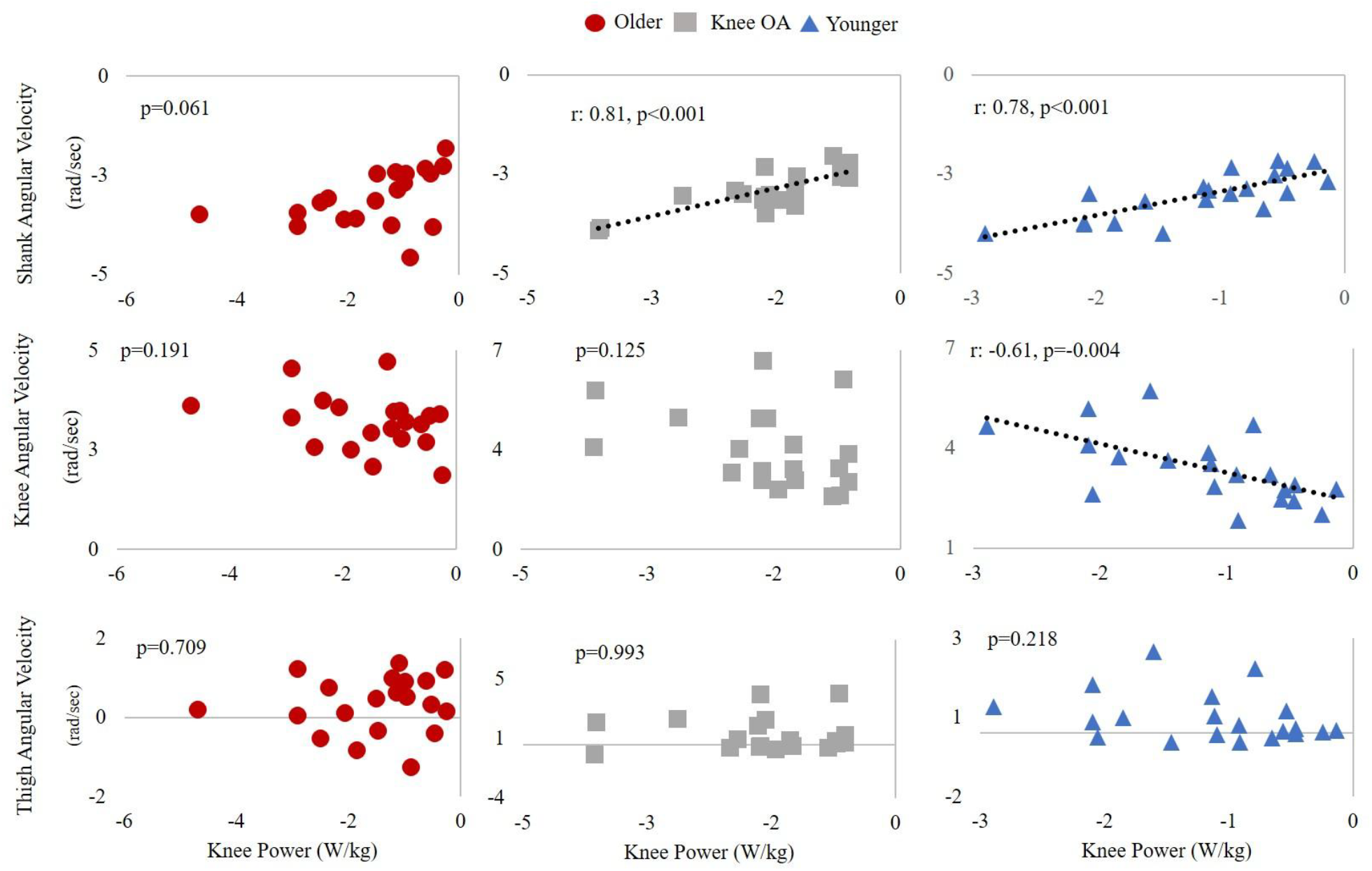
Early stance IMU-derived angular velocities plotted against early stance negative knee joint power for each group. p-values are shown for each comparison and trend lines and Pearson correlation coefficients are included where correlations were significant. All r values can be found in the text.

**Fig. 5.**
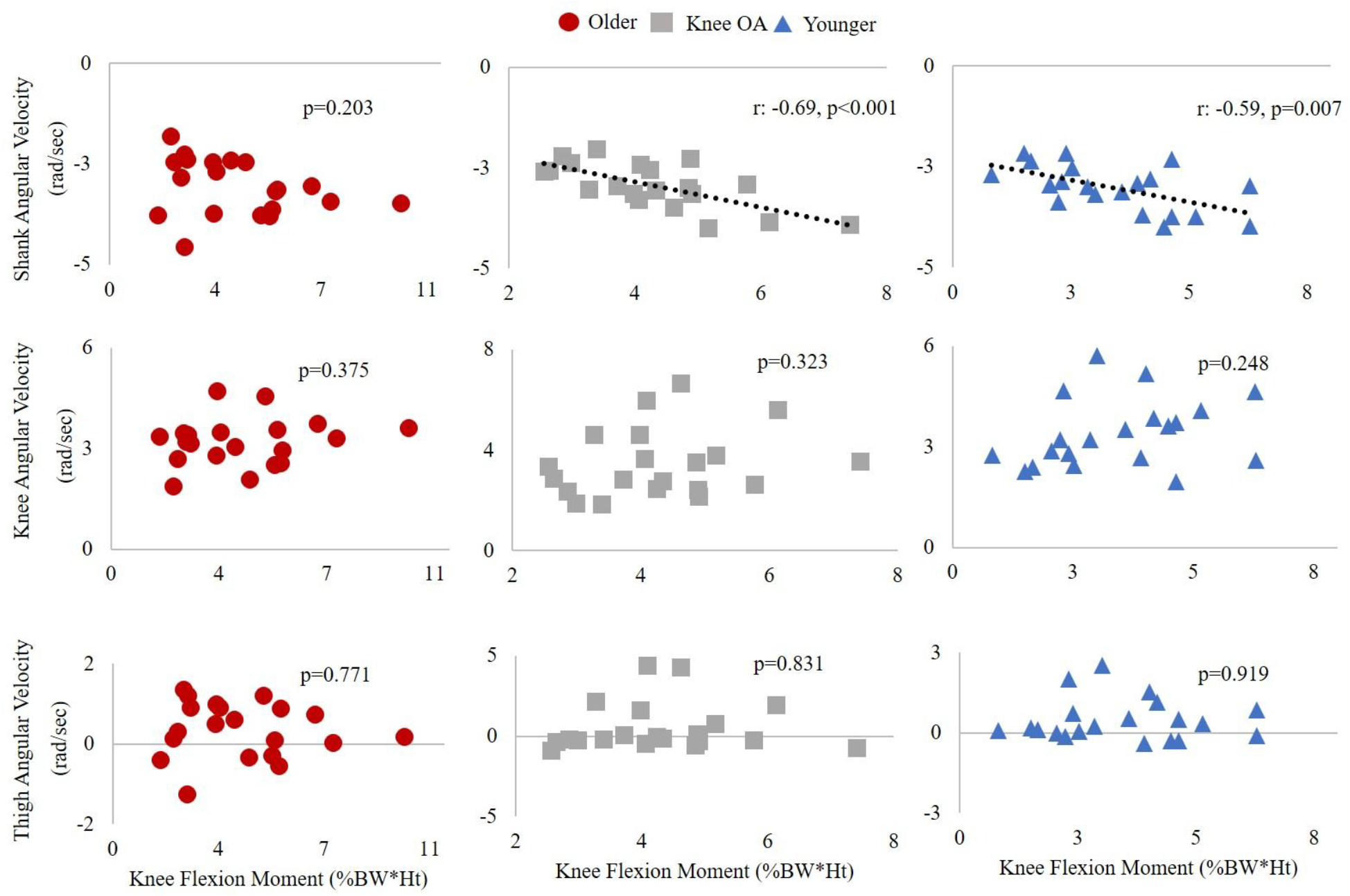
Early stance IMU-derived angular velocities plotted against early stance knee flexion moment for each group. p-values are shown for each comparison and trend lines and Pearson correlation coefficients are included where correlations were significant. All r values can be found in the text.

In the older adults, OA, and young groups, peak knee joint power did not correlate with thigh segment angular velocity (r=0.08, p=0.727; r=0.07, p=0.782; r=-0.28, p=0.239). Peak knee flexion moment correlated with shank segment angular velocity in the OA (r=-0.69, p<0.001) and young group (r=-0.59, p=0.007), but not in the healthy older group (r=-0.30, p=0.203). In the older adults, OA, and young groups peak knee flexion moment did not correlate with knee joint angular velocity (r=0.21, p=0.375; r=0.23, p=0.323; r=0.27, p=0.248) or with thigh segment angular velocity (r=-0.07, p=0.771; r=-0.05, p=0.831; r=-0.02, p=0.919).

## 5. Discussion

Traditionally, the measurement of joint kinetics has been limited to motion capture labs. This limits our ability to estimate joint loading and understand how kinetics affect health or change in the real world or in response to interventions. Our current study tested whether surrogate metrics of joint kinetics - IMU-derived joint and segment angular velocities - correlate with early stance negative knee joint powers and knee flexion moments. We found that IMU-derived shank and knee angular velocities correlated with early stance negative knee joint power and shank angular velocities correlated with knee flexion moments in a group of participants that included younger and older adults with and without knee OA. For within-group comparisons, shank angular velocity remained correlated with early stance negative knee joint power and knee joint moments for the healthy younger adults and adults with knee OA, but not for the healthy older adults. Our findings suggest that changes in knee joint kinetics during the early part of stance can be tracked using IMU-derived metrics.

The ability to track kinetics using IMUs in the lab could translate to tracking changes in kinetics in response to interventions in the clinic or in the real world. Previous studies have focused on directly predicting or estimating joint kinetics, while our study aimed to identify correlations between IMU-derived metrics and inverse dynamics measures. The presence of such correlations would suggest that changes in inverse dynamics could be detected, or tracked by, changes in IMU-derived surrogate measures. Our results support the ability to track changes in early stance knee joint kinetics with IMU data alone. This is in contrast to previous work that has estimated or predicted kinetics from IMU data using machine learning or musculoskeletal modeling. For example, Karatsidis et al. used Xsens sensors and methods to estimate body segment kinematics – variables that align with traditional motion capture outcomes – and then used musculoskeletal modeling and forward dynamics procedures to estimate kinetics [24]. Such procedures require body measurements and assumptions that go beyond IMU-alone measurements and can be computationally intensive. Other researchers have used machine learning algorithms to estimate ankle joint power [25] and knee flexion moments [10] from IMU data. The machine learning models in these studies used MoCap kinetic and kinematic training data, and the models’ sensitivity to their training data sets is unknown. In contrast, our work suggests that changes in kinetics could be tracked without participants visiting a lab and without collecting extensive training data.

Our findings suggest that the shank sensor alone may be sufficient to track negative knee joint power and knee flexion moments outside the lab. Using just one sensor for tracking joint kinetics would decrease patients’ burden of wearing multiple sensors and facilitate easier data processing and tracking of patient treatment progression. Our results are similar to previous work demonstrating that a single sensor may be sufficient to track ankle or hip joint powers [11]. The shank sensor data may provide a stronger correlation due to its better representation of rigid-body motion, while the thigh sensor would be subject to the larger soft tissue artifact of that segment.

In order to be confident that the current findings would translate to other settings or conditions, the correlation between IMU-derived data and early stance knee kinetics must be tested within an individual when their kinetics change. Stride length, gait speed and quadriceps muscle strength influence kinetics and can be manipulated relatively easily by dictating walking speed or cadence and by inducing muscle fatigue. Increased stride length and gait speed lead to larger knee flexion moments [17] and lower quadriceps muscle strength leads to reduced peak external knee flexion moments [26]. While our study collected data at two self-selected speeds, these speeds do not provide adequate data to model sufficient within-individual variation for testing correlations. Testing whether IMU-derived surrogate metrics of kinetics track with knee kinetics in a within-subjects design using a variety of conditions would strengthen the potential that this method would be useful in a clinical or intervention setting.

One limitation of our study is the potential that IMU angular velocities wouldn’t track changes in joint powers if a change in kinetics were due to something other than a change in angular velocities. Joint powers are the product of joint moments and angular velocity. If an individual’s change in knee joint power were due to a change in moments via either a change in ground reaction force or body posture rather than angular velocity, this IMU metric may not detect the change in kinetics.

## 6. Conclusion

This study demonstrates the potential of tracking negative knee joint powers and knee flexion moments with IMU-derived angular velocities as surrogates of kinetics. Additionally, we found that a single sensor on the shank is sufficient for tracking negative knee joint powers and knee flexion moments across groups. More work is needed to test the ability of angular velocity measured with IMUs to track changes in kinetics within individuals.

## Data Availability

All data produced in the present work are contained in the manuscript

